# Telemedicine and clinical care for Neglected Tropical Diseases in a post-pandemic world: where are we on the way to mitigate global inequities? A scoping review

**DOI:** 10.1101/2024.08.07.24311593

**Authors:** Fernanda G F Salvador, Claudia D Jardim, Mayumi D Wakimoto, Luis V Lapão, Henrique M C da Silveira, Cláudia M Valete

## Abstract

**Background:** Neglected Tropical Diseases (NTDs) represent a global public health problem with great socioeconomic impact. Using digital health technologies to provide medical care remotely can be an important strategy for reducing inequalities in access but is challenging in low-and middle-income settings and geographically isolated areas. The objective of the current review was to identify and summarize international evidence on the use of telemedicine for clinical care of patients with NTD around the world based on a scoping review protocol.

**Methodology/Principal Findings:** A total of 422 articles were extracted from the databases MEDLINE/PubMed, Web of Science and Scopus, and after remove 129 duplicates, 285 studies were excluded because they did not meet at least one of the eligibility criteria. A total of 8 articles were included for data collection, published between 2006 and 2023, half of them after 2021 (n=4). Fifty percent of the studies (n=4) were focused on dermatological diseases, like leprosy and leishmaniosis, and the other diseases found were dengue (n=2), trachoma (n=1) and cysticercosis (n=1). Most of telemedicine interventions in NTD took place in South America (n=5), with emphasis on Brazil (n=2). Studies that evaluated the accuracy of remote diagnosis demonstrated good effectiveness for leprosy, trachoma and complications of neurocysticercosis. There was a significant reduction in the need for specialized in-person medical consultations with telemedicine for the care of dengue fever and some dermatological NTDs; and an improvement in the quality of clinical monitoring of cutaneous leishmaniasis using mobile health was related.

**Conclusions/Significance:** This scoping review mapped the existing evidence on telemedicine applied at clinical care for NTD. Although we observed a small recent increase in academic research on the theme after COVID-19 pandemic, there is a gap in recommendations for the clinical management of NTDs through telemedicine, especially for synchronous approaches, revealing that this resource is still largely underutilized.

**Author Summary:** Neglected Tropical Diseases (NTDs) represent a global public health problem with great socioeconomic impact. Using digital health technologies to provide medical care remotely can be an important strategy for reducing inequalities. The objective of this scoping review was to identify and summarize international evidence on the use of telemedicine in the clinical care of patients with NTDs. A total of 8 articles were included for data collection as they met all our search criteria, half of them published after 2021 (n=4). Most of the studies took place in Latin America (n=5) and involved Primary Health Care teams (n=5). Studies that evaluated the accuracy of remote diagnosis demonstrated good effectiveness for leprosy, trachoma and complications of neurocysticercosis. There was a reduction in the need for specialized face-to-face medical consultations with telemedicine to treat dengue fever and some dermatological NTDs, and an improvement in the quality of clinical monitoring of cutaneous leishmaniasis through mobile health was reported. Although we have seen a recent small increase in academic research on the topic following the COVID-19 pandemic, there is a gap in recommendations for the clinical management of NTDs through telemedicine, especially for synchronous approaches, revealing that this resource is still largely underutilized.

## Introduction

Neglected Tropical Diseases (NTDs) are estimated to affect more than a billion people in poor and vulnerable regions of the world, but they occupy a disproportionate space in the priorities of the international agenda, reflected in low investments in research, therapeutic strategies and elimination/control actions [1, 2]. In addition to early mortality, disabilities caused by NTD perpetuate cycles of social exclusion due to unemployment and low education and are strongly related to socioeconomic conditions as irregular access to drinking water, basic sanitation and adequate housing. The control of these diseases is formally recognized as a goal of global action in the Sustainable Development Goals (SDGs) defined by the United Nations (UN) and the development of mechanisms to expand universal access to health is one of the main strategies recommended by the World Organization World Health Organization (WHO) to combat NTDs [1].

NTDs are prevalent in low- and middle-income countries (LMIC) that often have low coverage by health systems or irregular distribution of doctors between territories. Using digital health technologies to provide medical care remotely is an important strategy for reducing inequalities in access to healthcare [3, 4]. But, despite recent advances, this potential for remote assistance still appears to be scarcely explored [5]. Many NTDs are endemic in rural and remote areas and may require rapid guidance for specific clinical management (e.g. snakebite envenoming), with a great potential for the use of these strategies. In other situations, the low prevalence and/or high complexity of the pathology may require highly trained focal specialists for diagnosis, and teleconsultations with distant hospital centers can optimize timely care and early treatment of NTDs, reducing the time between the onset of symptoms and specialized clinical care.

The Covid-19 pandemic triggered a rapid expansion in the provision of remote care via telemedicine, making it an indispensable resource for the healthcare sector [6]. The growth of successful experiences in Digital Health can be observed especially in high-income countries, but the legal regulation of this practice is heterogeneous, and its incorporation has been slower in LMIC due to low internet connection and infrastructure barriers [7]. Health systems occupy different levels of technological maturity to incorporate innovations, and many poor countries will need sustainable financial support and international technical cooperation to implement initiatives in this field.

Telemedicine uses telecommunications and information systems to deliver remote healthcare services when patients and providers are separated by distance [3]. Different strategies can be useful during the therapeutic journey of NTD care via telemedicine, such as synchronous telecare between professionals and patients in real time, or the exchange of opinions between experts from distant services for difficult cases. Remotely sending data, such as images and videos, can also be a simple and useful way to carry out an asynchronous teleconsultation, and the choice of the most appropriate resources will depend on the clinical situation and local context [8].

Although new Artificial Intelligence paradigms are rapidly advancing in healthcare and machine learning algorithms are already a reality, there is still a long way to go to ensure reliability and safety in clinical decision-making using these digital tools alone [9]. The interface with health professionals is still fundamental in health care. Despite the recent wave of research into technological innovations, the literature about successful experiences of remote clinical care via telemedicine in contexts of populations affected by NTDs in the post-pandemic global scenario is unclear.

### Research questions and scope

The goal of this scoping review is to review and map existing literature on telemedicine applied to NTDs. The main research question that guided the review was: what is the current state of scientific evidence on the use of telemedicine in the clinical care of patients with NTDs across the world? We seek to summarize currently available evidence of the area and identify the main gaps in knowledge production.

The research also sought to characterize the contexts where telemedicine initiatives for NTDs were developed, and which are the main technologies used by them.

## Methods

This scoping review was conducted in accordance with the Joanna Briggs Institute (JBI) methodology for Scoping Reviews [10] and followed the methodological framework suggested by the Preferred Reporting Items for Systematic Reviews and Meta-Analyses for Scoping Review (PRISMA–ScR) [11], (S1 Table). The research protocol was registered on the Open Science Framework plataform (OSF) in March 2024 and can be accessed in: https://doi.org/10.17605/OSF.IO/XAKF5. The team was composed of academic researchers, clinical health professionals, and digital health specialists. Three databases and interfaces were chosen for their reliability and ease of searching with extensive MeSH terms: MEDLINE/PubMed, Web of Science and Scopus.

For the review we considered the list of 21 NTD and disease groups currently defined and prioritized by WHO [12], with the aim of delimiting the scope of this research. However, we recognize that local social determinants of health may constitute a broader range of infectious diseases of poverty in each region.

There are many conceptual definitions of telemedicine in literature. In this study we assume a WHO definition [8] which encompasses the following activities: 1. consultations between a remote person and a healthcare professional; 2. remote monitoring of the person’s health or diagnostic data by the provider; 3. transmission of medical data (e.g. images, notes and videos) to the healthcare provider; and 4. case management consultations between healthcare providers.

Finally, we report that, due to the characteristics of the regions studied, the team considered all available means of communication, from videoconferences to regular calls by telephone, for example, as acceptable telemedicine resources.

### Data sources and searching strategies

The keywords and their variations were defined by the working group based on literature review and sensitivity test of the terms in retrieving relevant articles. Combining these groups, we generate keywords lists for two categories (Fig. 1). An advanced search in the three selected databases were performed using Boolean operators to identify articles of interest published in the period between January 1, 2000, and February 26, 2024 (date of first retrieval).

**Figure 1.**
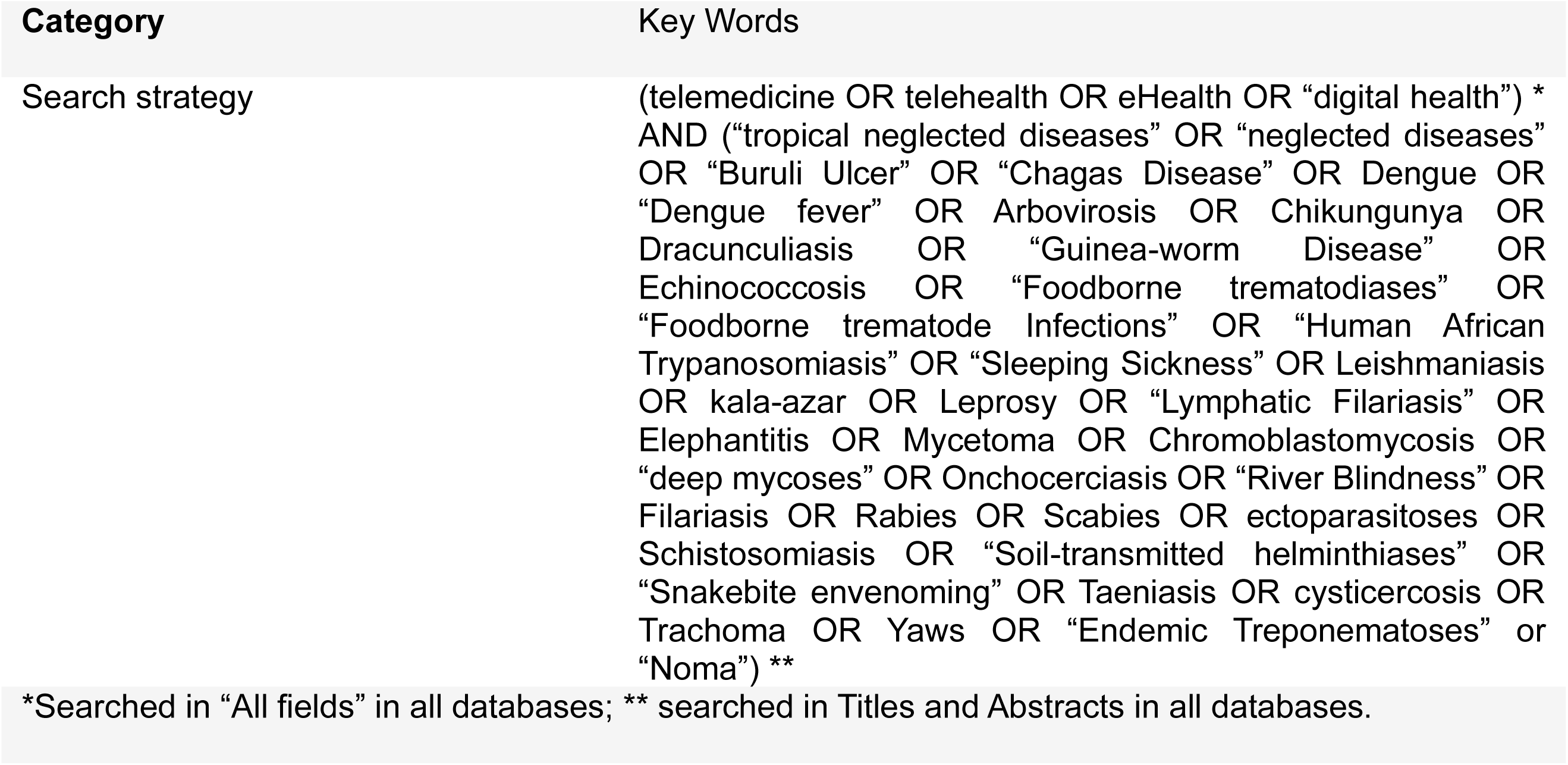
Search strategy.

Telehealth is a broad term for the use of Information and Communication Technologies to exchange health information remotely, which conceptually also refers to actions for purposes other than clinical disease care (e.g., health education or health management) [3]. This expression was also used in our primary search strategies, however only studies that carried out activities within the scope of telemedicine mentioned above were selected [8]. The same criteria were applied to studies retrieved by the terms Digital Health and eHealth (electronic health).

### Study selection and data extraction

The inclusion criteria were: 1. Practical experiences of using telemedicine for individual clinical care with direct interface between patients or the local provider and others healthcare professionals by distance; 2. Report at least one NTD from the WHO list as related to the telemedicine activity applied in the study; 3. Original peer-reviewed articles and other studies with different methodologies, namely: observational studies, implementation and intervention studies, pilot studies, qualitative, quantitative or mixed methods; and 4. Published between January 2000 and February 2024.

No language restrictions were applied. Systematic and scoping reviews, case studies, book chapters, editorials, grey literature and media sources were excluded.

Titles and abstracts were selected by two independent reviewers for evaluation according to the inclusion criteria using the Rayyan automation tool [13]. In the same way, the selected articles were read in full and classified. The steps were blinded to reduce bias. All disagreements that arose between the reviewers at each stage of the study selection process were resolved by a third reviewer.

A data extraction instrument for evidence synthesis was created in Microsoft Excel. We extracted data on the characteristics of the articles (e.g. authors, year, country of the studied population, main NTD/disease, study design, sample); contextual factors (e.g., main technology used, rural or urban area, type of care provider, team responsible for the telemedicine intervention) and main outcomes/final results.

The methodological quality of the studies was not thoroughly evaluated and no critical appraisal of individual sources of evidence was done.

## Results

### Eligible records

A total of 422 studies were initially retrieved. After excluding 129 duplicates and 250 titles and abstracts that did not meet at least one of the eligibility criteria, 43 articles were considered potentially relevant for full reading. A total of 8 articles were identified for inclusion in data extraction. The survey results were reported in full and presented in a PRISMA-ScR flow diagram (Fig. 2)

**Figure 2.**
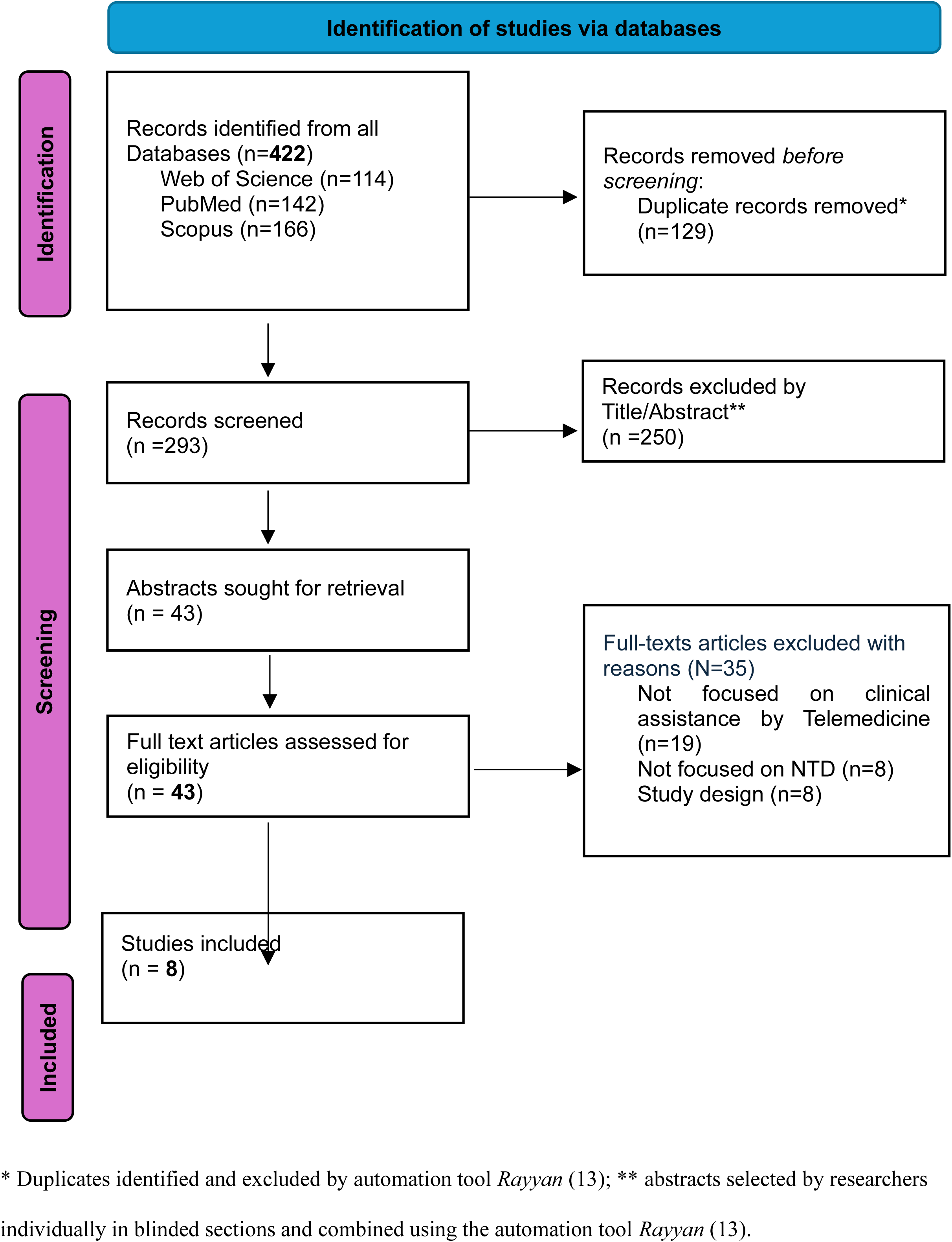
PRISMA-ScR chart.

### Study characteristics

The analysis of the final selection showed that 50% of the studies (N=4) were focused on dermatological diseases (14, 15, 16, 17): one specifically on leprosy (14) (N=1), one specifically on leishmaniasis (16) (N=1), and two in general dermatological diseases that included other dermatological NTDs in addition to leprosy and leishmaniasis, such as Buruli ulcer, scabies, yaws, tungiasis and filariasis (15,17). The remaining studies were related to the management of dengue (25%/N=2) (18, 19), cysticercosis (12.5%/N=1) (20) and trachoma (12.5%/N=1) (21). The majority of the 21 NTDs were not represented in the results (Table 1).

**Table 1.**
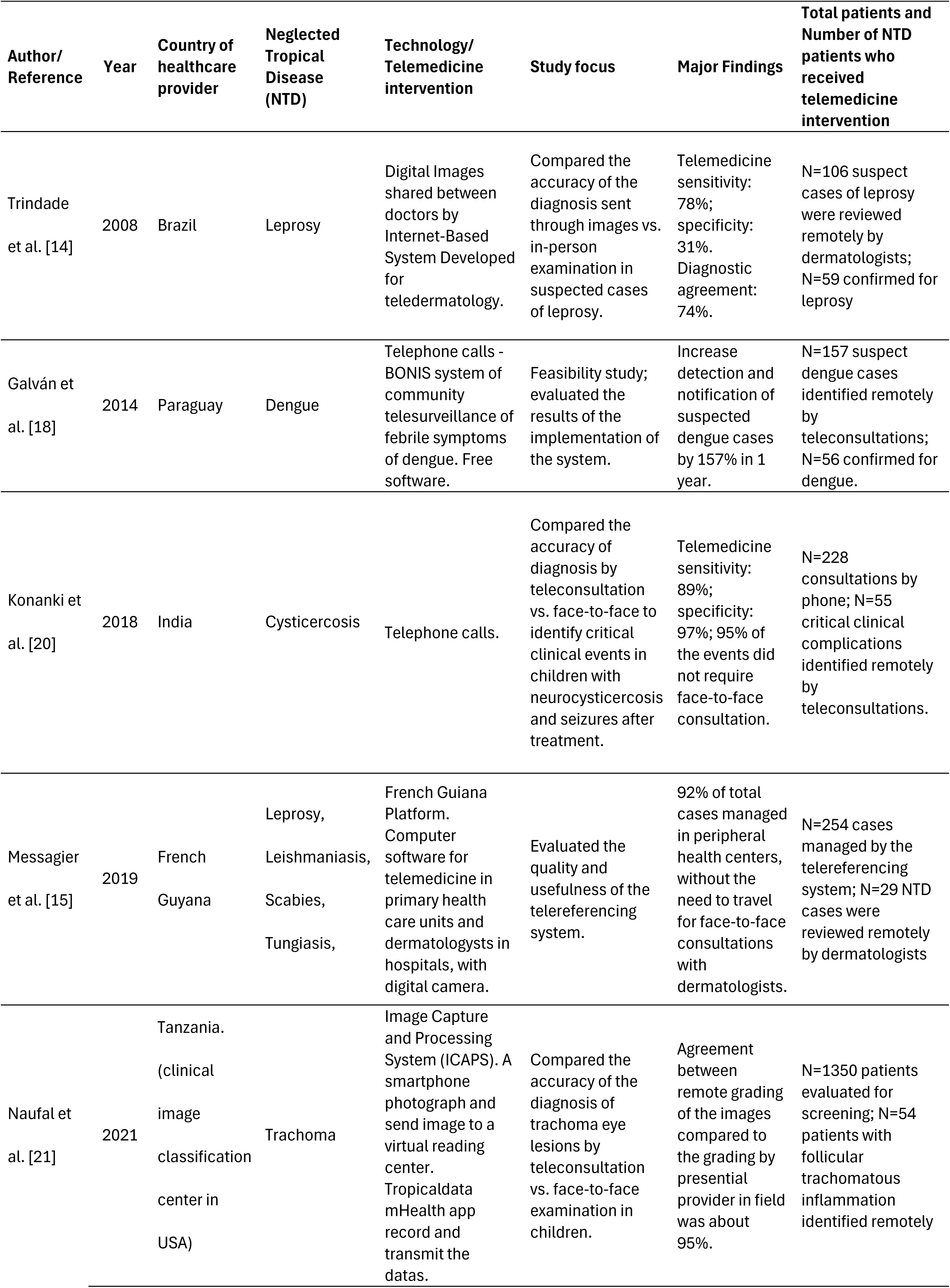
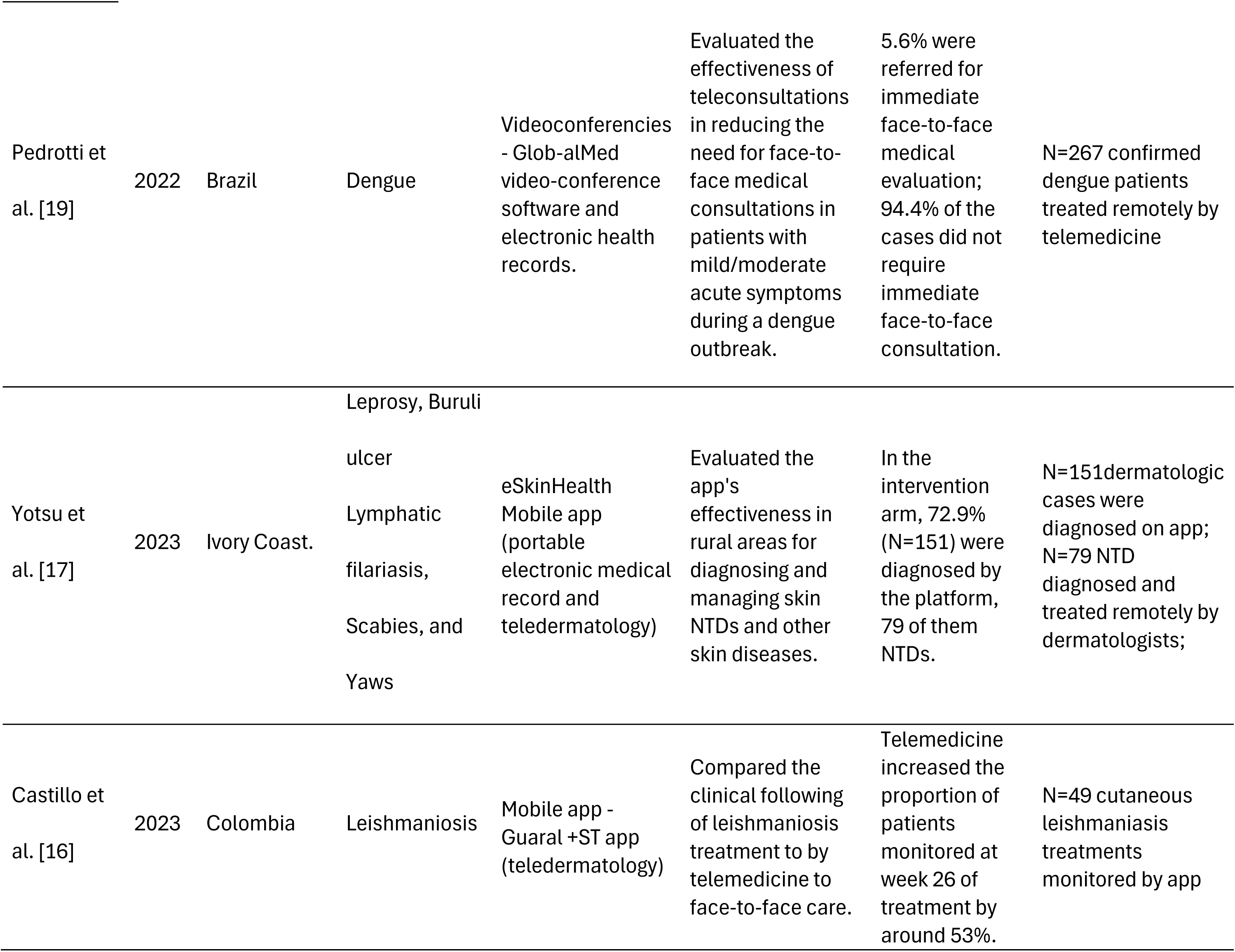
Characteristics of the studies included in the Scoping review.

The temporal analysis demonstrated that 50% of the studies (n=4) were published between 2021 and 2023, while the other 50% were published between 2008 and 2019, which points to an acceleration of studies after the Covid-19 pandemic, following the international trend (22). The length of the telemedicine intervention studies reviewed ranged from 2 to 18 months.

### Contexts

Regarding the geographic profile of the populations studied, 62,5% of the researches took place in South America (N=5), with emphasis on Brazil (N=2); 25%/ were conducted in Africa (N=2) and 12,5% in South Asia (N=1). Four studies were conducted in countries classified as upper-middle-income economies (23) (Brazil, Colombia, Paraguay), three studies in lower-middle income economies (India, Ivory Coast, Tanzania), and one in a high-income economy country (Guyana). No study was conducted in a country with a low-income economy.

A significant proportion of studies (62.5%, n=5) had telemedicine providers who were part of the local primary care team (São Paulo, Brazil; Asunción, Paraguay; Tumaco, Colombia; Cayenne, French Guyana; and Sinfra, Ivory Coast), and two of them utilized telemedicine to enable communication between the local team and the hospital (14, 15). One of the other studies was carried out only with staff from a tertiary hospital (New Delhi, India); one in a temporary emergency hospital due to a dengue outbreak (São Paulo, Brazil); and one worked in conjunction with a district-level community prevalence research team (Chamwino, Tanzania), which is still the only study where one arm of the clinical assessment was carried out in a different country (United States).

Finally, half of the 8 studies (50%, N=4) developed at least one of the arms in remote and/or rural areas (15, 16, 17, 21), three in urban settings (14, 18, 19) and in one it was not described (20).

### Telemedicine interventions and outcomes

Table 2 summarizes the characteristics of the telemedicine technologies studied. Only three studies investigated real-time synchronous interventions of consultations between remote patients and healthcare providers, two of which were via ordinary telephone calls (18, 20) and only one via videoconference (19). In all five other articles, telemedicine was carried out with asynchronous transmission of medical data (e.g. images, descriptive clinical notes), involving teleconsultations for case management between healthcare professionals located in different services (14, 15, 16, 17, 21).

**Table 2.** Thematic framework - characterization of the telemedicine interventions and contexts Telemedicine intervention.

|  | N | References |
| --- | --- | --- |
| **Telemedicine intervention** |  |  |
| Consultations between remote person and healthcare provider - real-time /synchronous telemedicine | 3 | [18], [19], [20] |
| Transmission of medical data (e.g. images, notes) mediated by distant healthcare providers - asynchronous telemedicine | 5 | [14], [15], [16], [17], [21] |
| **Communication technology** |  |  |
| Telephone | 2 | [18], [20] |
| Videoconference | 1 | [19] |
| Mobile application | 3 | [16], [17], [21] |
| Data/images submission platform | 2 | [14], [15] |
| **Clinical applicability** |  |  |
| Telemedicine to ensure patient access to diagnostic | 6 | [14], [15], [17], [18], [20], [21] |
| Telemedicine to ensure patient access to treatment | 1 | [19] |
| Telemedicine to monitor patient treatment | 1 | [16] |
| **Telemedicine providers involved** |  |  |
| Primary Health Care team | 5 | [14], [15], [16], [17], [18] |
| Hospital team | 4 | [14], [15], [19], [20] |
| Other | 1 | [21] |
| **Location where the study population resides** |  |  |
| Rural or/and remote | 4 | [15], [16], [17], [21] |
| Urban | 3 | [14], [18], [19] |
| Not mentioned | 1 | [20] |
| Latin America | 5 | [14], [15], [16], [18], [19] |
| Africa | 2 | [17], [21] |
| Asia | 1 | [20] |
| Upper-middle-income economy | 4 | [14], [16], [18], [19] |
| Lower-middle-income economy | 3 | [17], [20], [21] |
| High income country | 1 | [15] |
| **Disease studied** |  |  |
| Leprosy | 3 | [14], [15], [17] |
| Leishmaniosis | 2 | [15], [16] |
| Dengue | 2 | [18], [19] |
| Scabies | 2 | [15], [17] |
| Cysticercosis | 1 | [20] |
| Trachoma | 1 | [21] |
| Tungiasis | 1 | [15] |
| Buruli ulcer, Lymphatic filariasis, Yaws | 1 | [17] |

Regarding the telemedicine software used, two studies worked with specific telemedicine platforms for sending clinical data and images (14, 15) and one study tested a software for medical videoconferencing (19), while the use of mobile applications for remote care of NTDs occurred in three studies (16,17,21).

Most studies (75%, n=6) sought to evaluate the applicability of remote NTD telediagnosis, but only three of them (37,5%) compared the effectiveness/accuracy of remote diagnosis via telemedicine with in-person medical care as a gold standard (14, 20, 21), obtaining favorable results for the use of teleconsultation. Other primary results on the use of telemedicine found in the studies were: a significant reduction in the need for medical in-person specialized consultations (37,5%/n=3) (15, 19, 20); increased disease detection capacity (25%/n=2) (17, 18), and improved quality of clinical follow up by healthcare providers using a mobile app (12,5%/n=1) (16).

## Discussion

This scoping review allowed the general mapping of available scientific production related to NTD care via telemedicine. The majority of the 21 NTDs were not represented in the results. The study demonstrated that there is a gap in recommendations for the clinical management of acute and chronic NTDs through telemedicine, revealing that this resource is still largely underutilized for the care of this group of infectious diseases of poverty. By way of comparison with a single other infectious disease, a bibliometric study published in May 2022 that investigated the application of Telemedicine in the care of COVID-19 retrieved 5,224 research papers published up to that time (24). A number 12,37 times greater than what we found for the 21 NTDs together until 2024.

Studies that evaluated the accuracy of remote diagnosis demonstrated good effectiveness for leprosy, trachoma and complications of neurocysticercosis. There was also a significant reduction in the need for in-person specialized medical consultations when telemedicine alternatives were implemented for the care of dengue fever and some dermatological NTDs, in addition to an improvement in the quality of clinical monitoring of cutaneous leishmaniasis using mobile health technologies. Low-income countries appear to remain excluded from advances in telemedicine for NTD, suggesting that the challenges of implementing remote care services in extreme poverty contexts are a relevant gap and an imminent challenge.

### Remote care for Neglected Tropical Diseases

Cutaneous NTDs stood out both for their applicability and level of evidence in remote clinical monitoring. The main findings in this review around teledermatology were already expected and corroborate the evidence on the previous consolidation of this specialty in the international telehealth scenario, including NTDs (25, 26). The ease of managing clinical parameters in dermatology through photographs and videos, increasingly accessible on modern devices, seems to be a facilitator. The use of mobile phones for healthcare (mobile health, currently called mHealth) is a growing market that was also shown to be more focused on neglected skin diseases than other NTDs in our study results. It is important to highlight, however, that there are also limitations that make substitutive and exclusive longitudinal follow-up via telemedicine impossible for some diseases. For cutaneous leishmaniasis, for example, the mucosal lesions would be difficult to verify through photographs and would require in-person visits to the service.

Dengue is a seasonal NTD transmitted by mosquitoes with high morbidity that can overwhelm emergency services. The use of telemedicine in dengue outbreaks has emerged as an effective strategy for early diagnosis and medical assessment of low-to-moderate risk cases, expanding case identification and preventing progression to severity. However, the urgency of the global epidemiological scenario of the disease (27) suggests that this resource could be more often used by health systems in order to avoid preventable deaths in areas with limited health service coverage. Digital health consultations combined with point-of-care diagnostic methods, such as rapid tests, is a window of opportunity to expand the differential diagnosis of acute febrile illnesses in the large endemic regions of Latin America and Southeast Asia and may also be useful in African regions where there is the challenge of differential diagnosis with malaria.

Trachoma is an infectious eye disease and an important cause of blindness in poor, rural and remote areas of the world. In our review we observed that the integration between screening programs and remote diagnosis by telemedicine can be a feasible advance in its elimination plan. One study uses a mHealth application suggesting an advance incorporation of mobile health resources in trachoma care (21). However, no telehealth actions focused on preventive chemotherapy for trachoma in endemic communities were identified in our review. The development of integrated preventive chemotherapy telemonitoring actions could be beneficial to support community strategies for the control and elimination of other NTDs, such as onchocerciasis, lymphatic filariasis, schistosomiasis and soil-transmitted helminthiasis in isolated communities at risk.

Clinical monitoring by telephone to track seizures in patients with neurocysticercosis proved to be a relevant strategy for remote monitoring of this chronic disease, avoiding unnecessary trips to clinical follow-up, without the need for sophisticated technology (20). For other long-term NTDs typical of remote rural areas, such as Chagas Disease and Human African Trypanosomiasis, no results were recovered, although it was expected that continuous chronic telemonitoring of these patients could have good applicability, as it would avoid unnecessary trips to distant specialized centers for clinical monitoring of complications. Despite the emphasis that zoonotic diseases have received around the world after the Covid-19 pandemic, and the advancement of the international debate on One Health approaches, the clinical management of some zoonotic NTDs seem to out of sight by the global digital health agenda, with technological advances more concentrated in sectors like vectors surveillance (28, 29) or digital microbiology (30).

### Telemedicine scenarios

Most interventions took place in Latin America. We identified a shortage of studies in African countries and the lack of innovative health experiences aimed at this region is a permanent challenge in controlling NTDs. Although the African continent accounts for around 40% of the global NTD burden (31), its broadband internet penetration rate was still around 33% in 2020 (32). Applications that work offline and do not require constant Internet can be part of the solution, but the challenge of providing access to person-centered services of remote healthcare especially in the sub-Saharan region requires complex multisectoral efforts and local engagement. The shortage of skilled health workforce in this region is a critical problem for addressing prevalent diseases, and consultations with specialists located in distant centers may be a good alternative in supporting infectious disease care (33). Furthermore, investment in public health systems with broad coverage can contribute to decentralized care in communities affected by NTDs, but also requires local governance policies committed to this agenda.

NTDs are closely related to the territory’s environmental health conditions, and Telemedicine applied at the level of primary health care, as seen in most of the studies, has the potential to expand digital community strategies for local surveillance and better epidemiological control. Improving the management of clinical communication between different levels of care (e.g. primary care centers and tertiary hospitals) through integrated Telemedicine platforms can also bring potential benefits to continuity, comprehensiveness and clinical coordination, considered internationally as important pillars for strengthening health systems centered on primary care (34). These results are also important to strengthen joint action between general practitioners and focal specialists through telemedicine in the management of patients with NTD.

### The implementation of telemedicine

The scarcity of results on long-term telemedicine implementations limits the understanding of the real barriers to its incorporation into services. As most of the studies identified in this review consisted of proofs of concept or pilot tests, it would be important to investigate which initiatives remained in operation, to assess the factors that may be associated with their success and sustainability in real-world settings over time. The integration of digital health systems into care networks emerges as a challenge, especially in health systems with few resources, as their effectiveness depends on local infrastructure, internet accessibility and connection quality, in addition to disruptive changes in the existing healthcare routine. However, the real impact of this operation on the long-term routine of providers was not explored in depth in most of the selected studies.

On the way to mitigate global inequities in access to internet and communication/information technologies in health, the implementation of telemedicine on a large scale requires complex efforts from several areas for its incorporation into health services: legal rules; systems interoperability; person-centered clinical telemedicine protocols; specific training for health providers; and promotion of patients’ digital literacy - which can be difficult for populations with low socioeconomic status. Another aspect that should be considered in telemedicine aimed at NTD is digital security and ethical regulation practices. Concern about ethical care when sending sensitive health data virtually is mandatory in the development of safe telemedicine tools in poor countries, that sometimes have limited legislation to guarantee the data protection of vulnerable populations.

Most of the initiatives presented in the studies occurred asynchronously. Only one study presented a structured telemedicine platform that offered teleconsultation via videoconference in real time between patients and doctors, but it did not eliminate the need for the patient to travel to the health service, since the medical teleconsultation took place after an in-person assessment by nursing staff. Henceforth, the use of mobile applications for remote care for NTDs emerges as a promising resource (16,17,21).

The absence of local validated clinical protocols for remote video consultations that replace face-to-face care could also make clinical research in this area still fragile from an ethical-legal perspective, especially in acute complications with potential for severity. Therefore, the development of guidelines for remote care for NTDs, standardizing safe remote care, also emerged as an important gap to be filled, since nothing in this regard was mentioned in the studies.

### Limitations

Publication bias is a limitation of this work, as we only considered articles published in journals and the search did not cover grey literature. To overcome this problem we used broader search strategies to map local experiences originating especially in low-income settings.

As NTD constitute a clinically diverse group of diseases, with different durations, stages of evolution and possible complications, in-depth comparative variables for intervention approaches are scarce. However, we highlight the main common characteristics and interventions aspects.

The group of neglected fungal diseases is not specified in the WHO list, described as “mycetoma, chromoblastomycosis and other deep mycoses” (12), making it difficult to include each of these diseases in the search strategy (Fig. 1), which may contribute to selection bias. For example, although paracoccidiomycosis and histoplasmosis are relevant problems in the Americas, we did not find consistent evidence to support the nominal inclusion of these pathologies in our review search strategy (35,36).

## Conclusion

Telemedicine applied to NTDs is not a new field, but the results of this review demonstrate that the use is limited compared to the global magnitude of NTDs. Although there is a recent increase in academic production on the topic after the COVID-19 pandemic, the evidence is restricted to a small group of diseases and the technologies used were, in most cases, asynchronous.

Telemedicine has emerged globally as a potentially promising resource for reducing disparities in access to healthcare services. Although its popularity has grown following the pandemic experience, the research gap regarding its effectiveness in the clinical treatment of NTDs is still large. The limited data available from these studies suggests that telemedicine is feasible compared to conventional care, but further in-depth studies are needed to compare in-person and remote consultations and to investigate the level of quality of current evidence and the benefits of this modality. Cost-effectiveness results are also needed to support decision-makers, especially in poor healthcare settings.

## Data Availability

The research protocol where the data used in this submission can be accessed on the Open Science Framework plataform (OSF) IN: https://doi.org/10.17605/OSF.IO/XAKF5. The funder had no role in study design, data collection and analysis, decision to publish, or preparation of the manuscript.

## Information support

### Funding

The main author of the article receives a doctoral scholarship from the Foundation for Science and Technology (FCT Portugal – www. fct. pt) under DOI registration 10.54499/UI/BD/151068/2021 (https://doi.org/10.54499/UI/BD/151068/2021)

The funder had no role in study design, data collection and analysis, decision to publish, or preparation of the manuscript.

### Competing interests

The authors declare that there are no conflicts of interest related to this article.

### Contributors

CV conceptualized the project and study design and was responsible for resolving divergent synthesis decisions. FS and CJ conducted the search and screening of literature and data extraction. FS led the theme analysis and writing of the manuscript. MW, LL and HS supported methodological and writing review of the manuscript.

## Supporting information

### Supplement 1

**Table S1.**
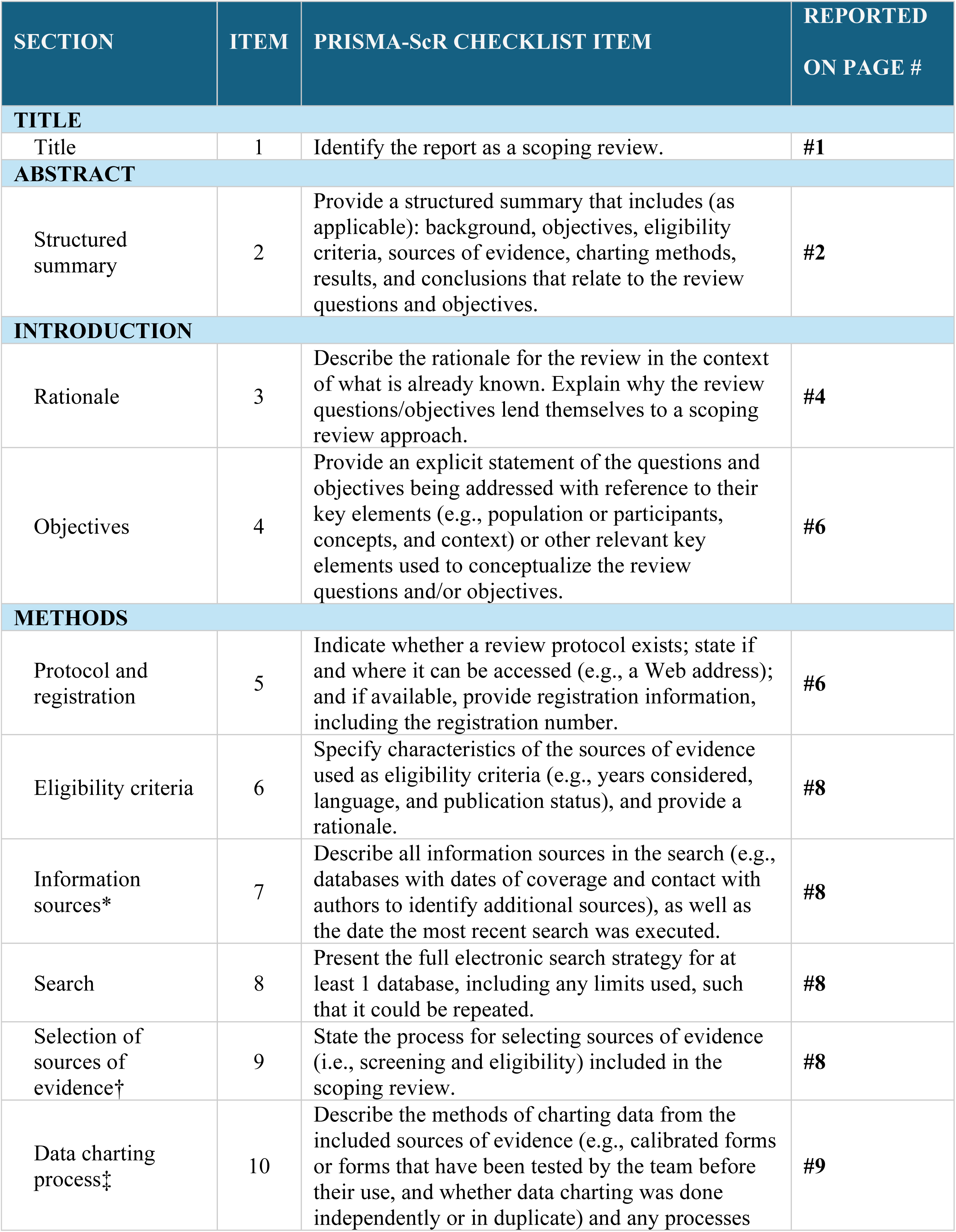
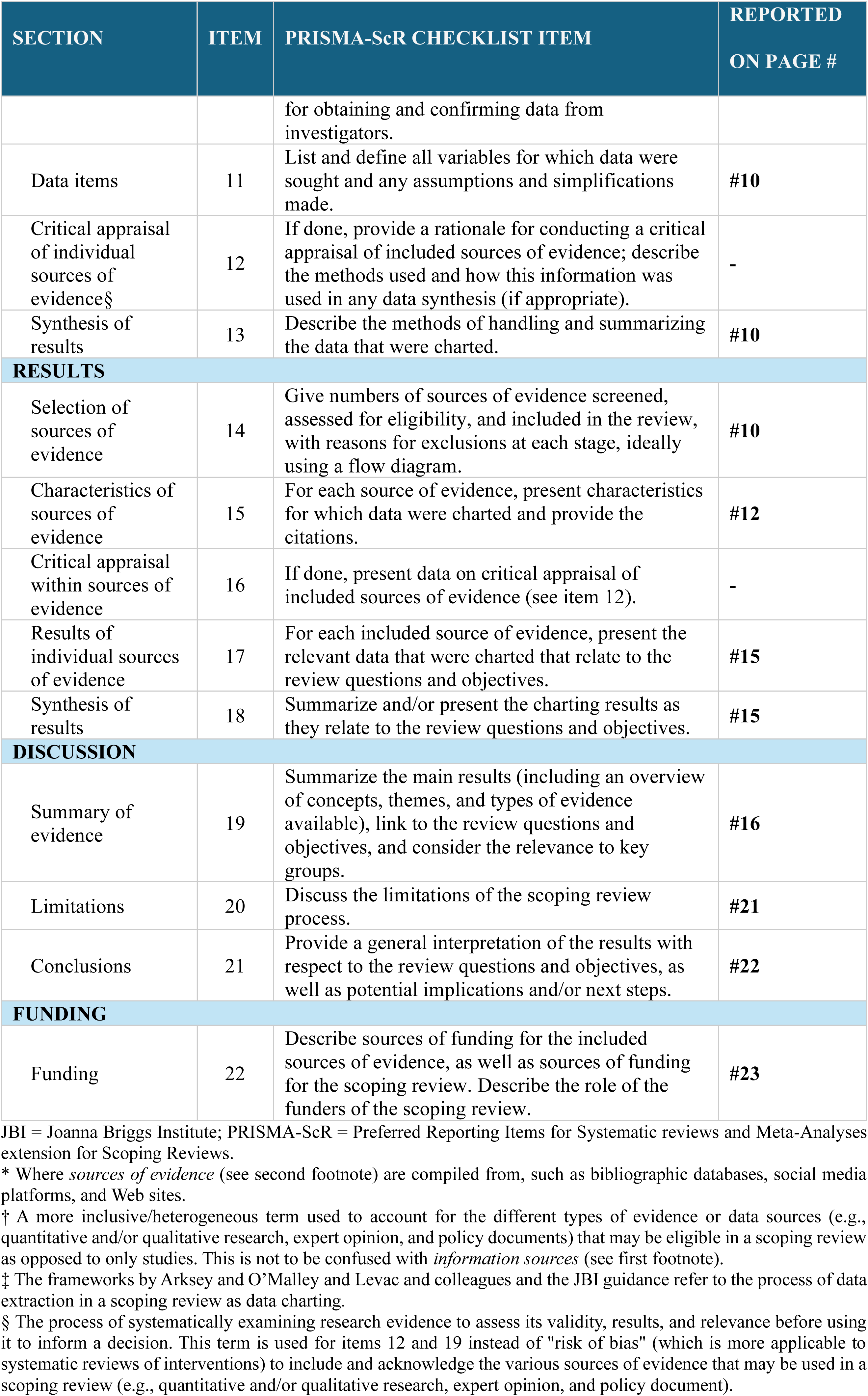
Preferred Reporting Items for Systematic reviews and Meta-Analyses extension for Scoping Reviews (PRISMA-ScR) Checklist applied for the review with respective corresponding pages.

## Notes

### Competing Interest Statement

The authors have declared no competing interest.

### Clinical Protocols

https://doi.org/10.17605/OSF.IO/XAKF5

### Funding Statement

Yes

### Author Declarations

This is a scoping review and therefore does not require ethical approval.

